## Supplementary figures and images for "Machine learning-guided deconvolution of plasma protein levels"

### Fig. EV1

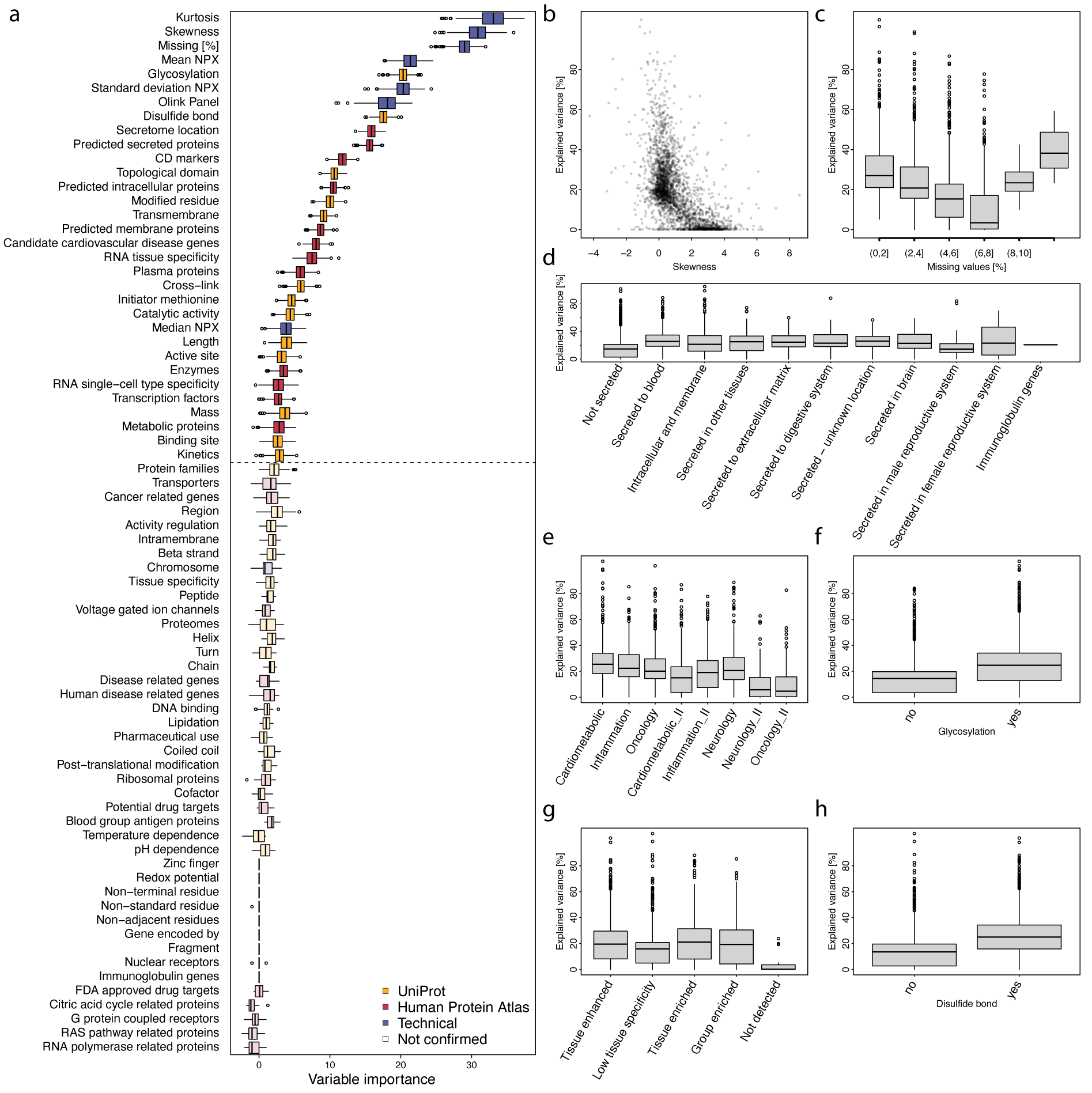

### Fig. EV2

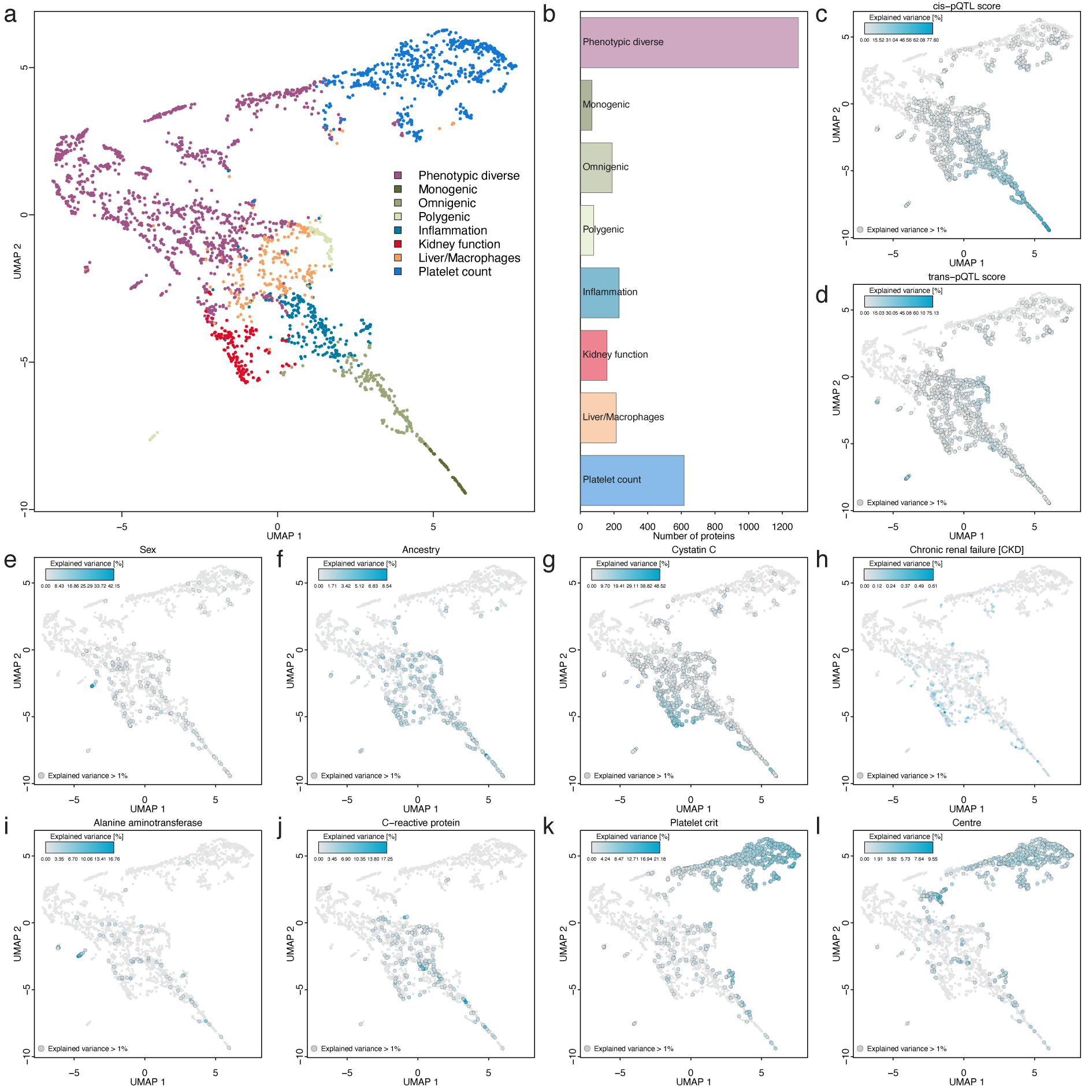

### FIG. EV3

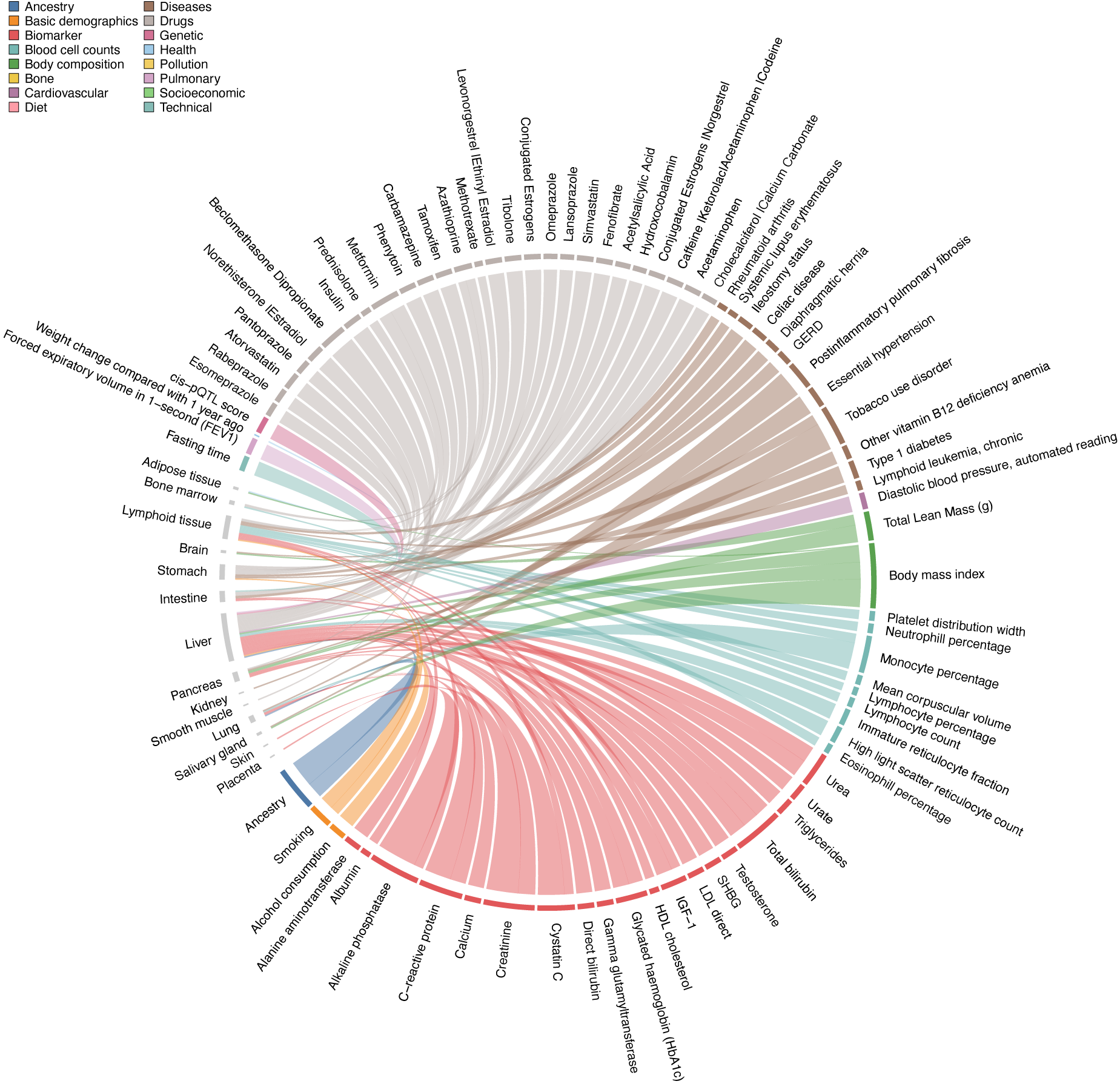

### Fig. EV4

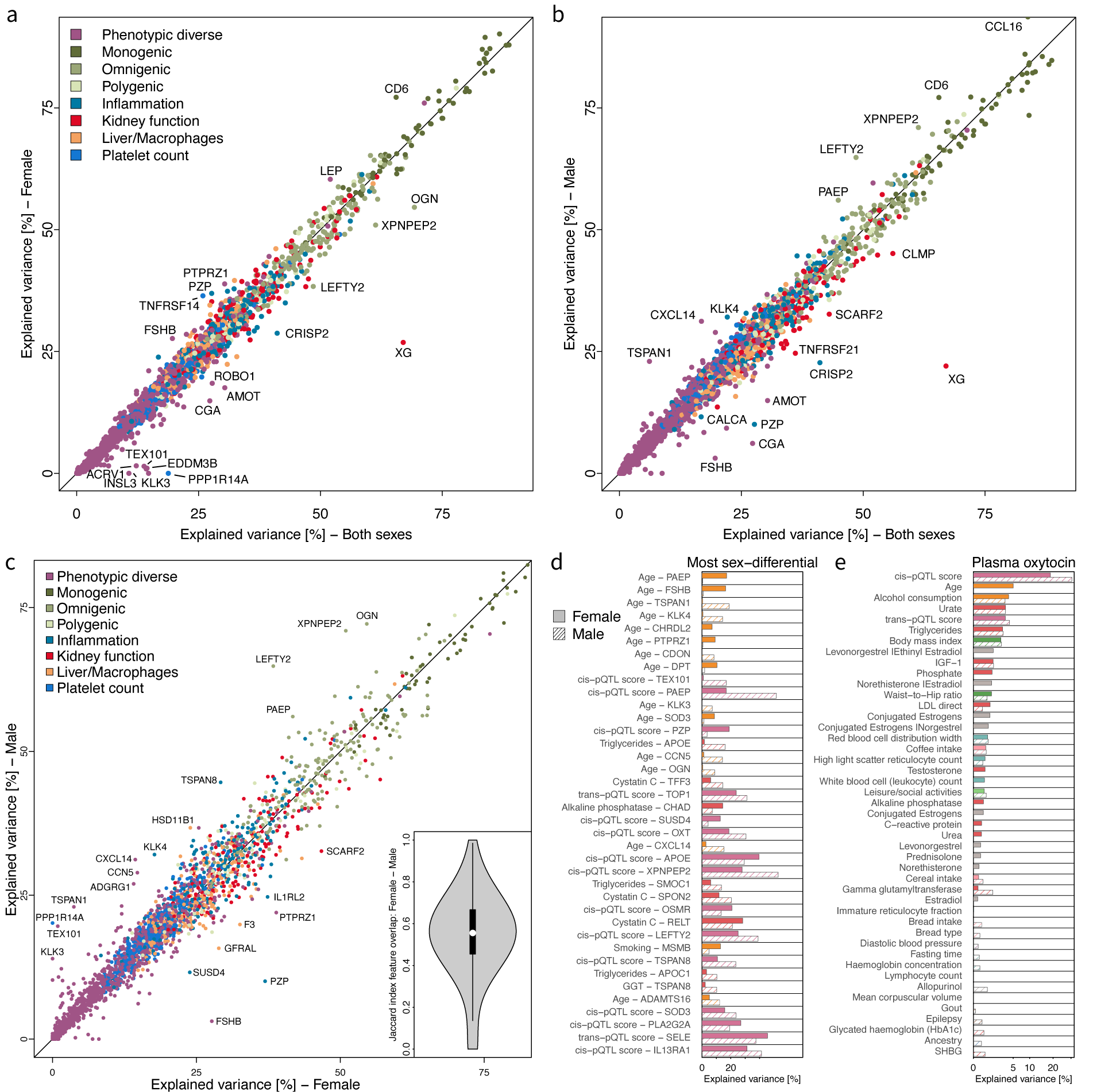

### Fig. EV5

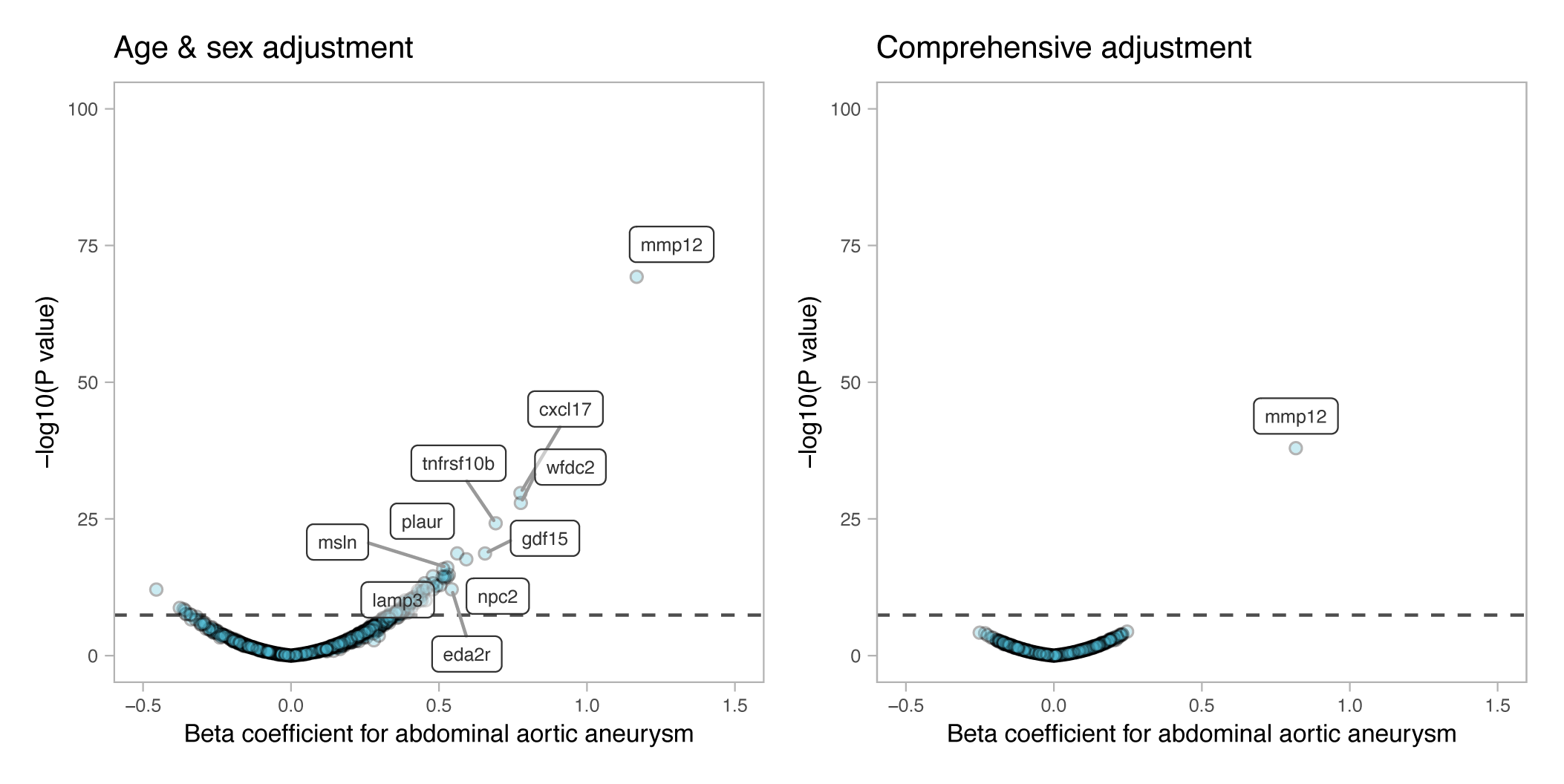

### Fig. EV6

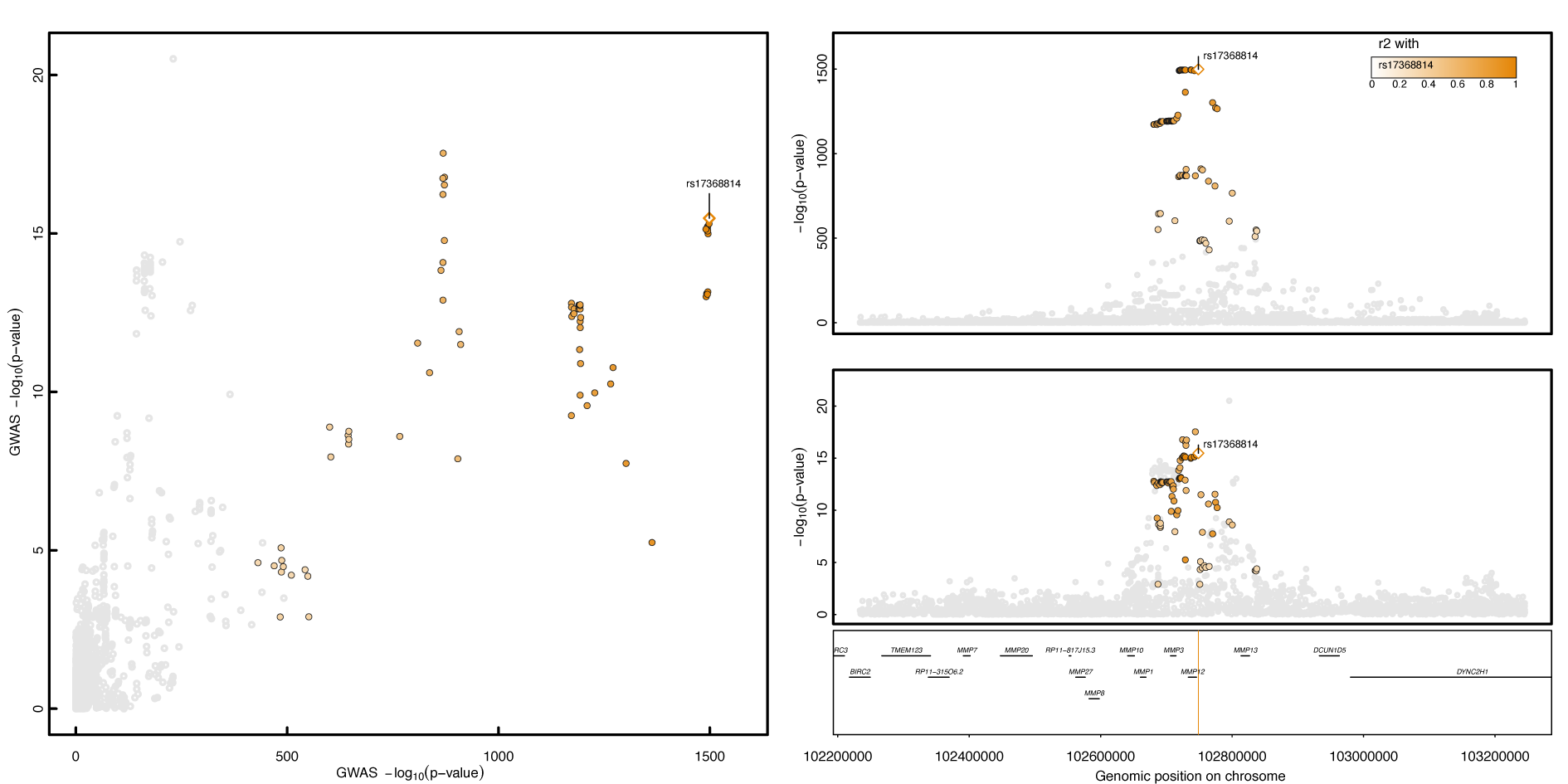

### Fig. EV7

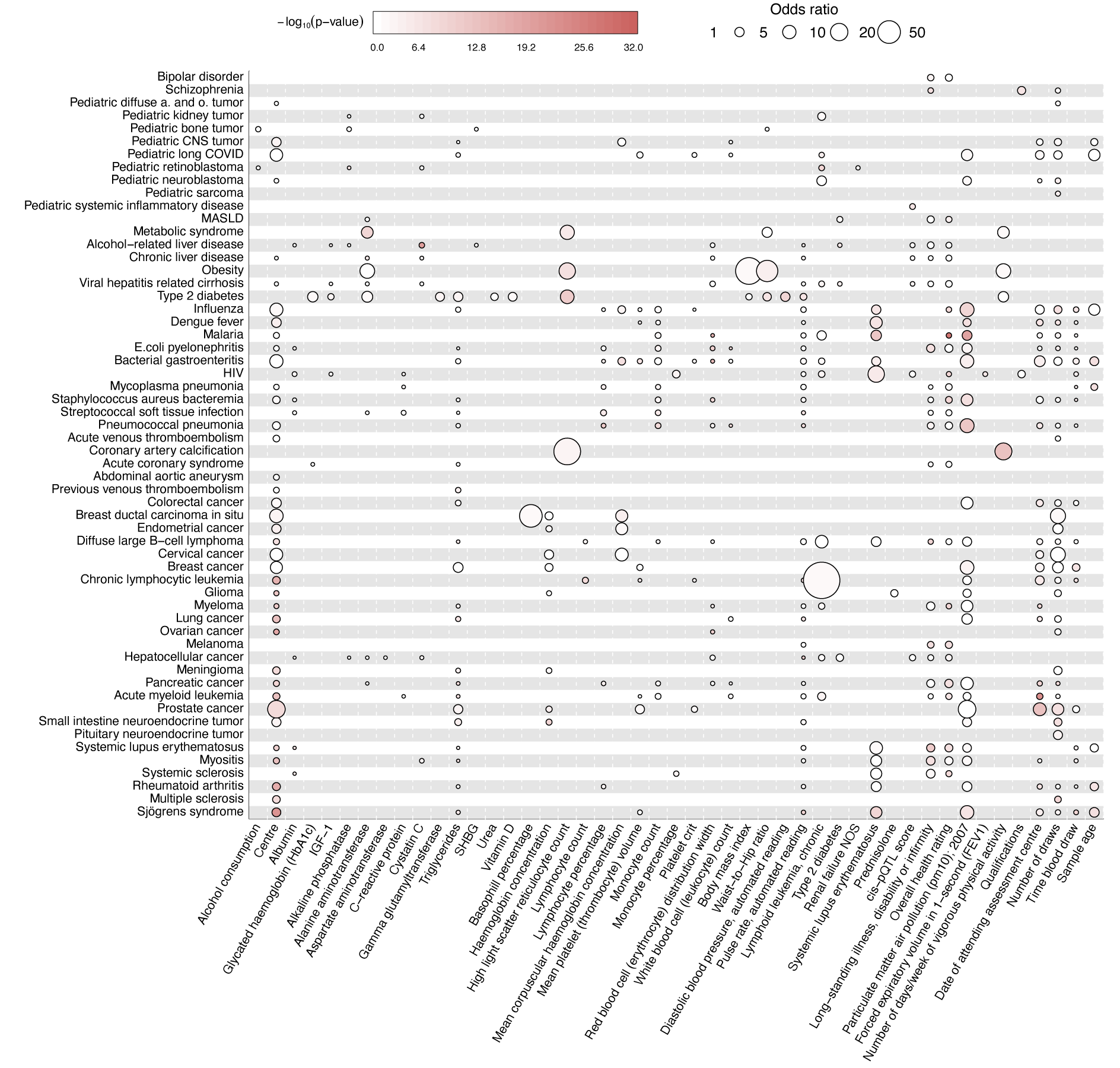
